# Unjustified Poisson assumptions lead to overconfident estimates of the effective reproductive number

**DOI:** 10.1101/2025.07.31.25332479

**Authors:** Barbora Němcová, Isaac H. Goldstein, Jessalyn Sebastian, Volodymyr M. Minin, Johannes Bracher

## Abstract

Time-varying effective reproductive numbers of infectious diseases are commonly estimated using renewal equation models. In the widely applied `R` package `EpiEstim` and various related tools, this approach is combined with a Poisson distributional assumption. This has been criticized on various occasions, mostly on grounds of general model realism or a desire to estimate overdispersion parameters. Here we argue that an important issue arising from the Poisson assumption is that inference about the effective reproductive number becomes overconfident in presence of overdispersion. By how much standard errors are underestimated follows in a straightforward manner from theory on generalized linear models. We therefore recommend to replace the Poisson assumption by quasi-Poisson or negative binomial extensions, and contrast their respective properties. We illustrate our arguments in detailed simulation studies and three examples of case studies of Ebola, pandemic influenza and COVID-19.

## 1 Introduction

The effective reproductive number, defined as “the expected number of new infections caused by an infectious individual in a population where some individuals may no longer be susceptible” (Gostic et al., 2020), is a key quantity characterizing disease outbreak dynamics. It is commonly estimated using renewal equation models, often combined with Poisson distributional assumptions. This approach was popularized by the `R` package `EpiEstim` (Cori et al., 2013), which due to its broad applicability, ease of use, and thorough documentation has become a standard tool (Nash et al., 2022). It has also been integrated in numerous methodological extensions (e.g., Brizzi et al. 2022, Green et al. 2022, Nash et al. 2023) and alternative `R` packages including `EpiInvert` (Alvarez et al., 2021), `ern` (Champredon et al., 2024), `estimateR` (Scire et al., 2023) and most recently `Rt.GLM` (Nouvellet, 2025). In this paper we are concerned with the uncertainty surrounding estimated reproductive numbers based on the renewal equation and a Poisson assumption for the observed incidence. We demonstrate that the associated standard errors are underestimated in the presence of overdispersion, a common feature of infectious disease surveillance data. Especially when incidence is high, uncertainty intervals based on Poisson assumptions get vanishingly narrow, overstating the certainty of the inference. While this has been pointed out empirically (Brockhaus et al., 2023; Goldstein et al., 2023), we provide a concise summary of relevant statistical properties. We moreover contrast several methods to account for overdispersion, which turn out to impact not only estimated standard errors, but also point estimates.

The remainder of the paper is structured as follows. Section 2 introduces the classic Poisson renewal equation model and outlines challenges arising from overdispersion, along with possible model extensions. Section 3 illustrates our arguments using simulations and case studies on COVID-19, Ebola and pandemic influenza. Section 4 concludes with a discussion.

## 2 Methods

### 2.1 The Poisson renewal equation model

The *instantaneous* reproductive number is often denoted by *R*_e_ or *R*_*t*_, but as in `EpiEstim` and related approaches it is assumed constant over an estimation window of *T* days, we simply write *R*. Daily or weekly incidence counts *X*_*t*_, *t* = 1, …, *T* are linked to *R* via the renewal equation (Fraser, 2007) and a Poisson assumption,

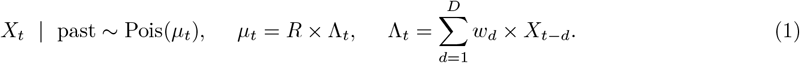

Here, *µ*_*t*_ is the expectation of the Poisson distribution. The discrete-time serial interval distribution, expressed by probabilities *w*_1_, …, *w*_*D*_, is considered known. The Poisson assumption is chosen as it makes Bayesian inference straightforward without the need for Monte Carlo methods (Cori et al., 2013).

Throughout the paper, we refer to *w*_1_, …, *w*_*D*_ as the distribution of the serial interval (the time between the onset of symptoms in a pair of consecutive cases) rather than the generation time (the time between two consecutive infections), as the exact times of infection are typically unobserved. It is known that using the serial interval distribution, which has a larger variance than the generation time distribution, introduces some light bias in the estimation of reproductive numbers (Britton and Scalia Tomba, 2019). However, this has become standard practice (Cori et al., 2013), and we do not explore this aspect in detail.

Equation (1) can be read as a Poisson regression with an identity link, no intercept and Λ_*t*_ as the sole covariate (Goldstein et al., 2023; Brockhaus et al., 2023). The properties of the resulting estimates can hence be studied in the framework of generalized linear regression (Fahrmeir et al., 2009). This is most straightforward for maximum likelihood (ML) estimates; the results, however, also translate to the Bayesian approach by Cori et al. (2013), which behaves similarly unless incidence is very low (Supplement B.1.2). The ML estimator takes the intuitive form

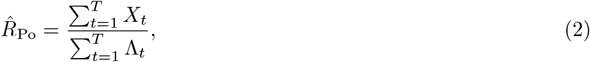

where the index “Po” indicates the underlying distributional assumption. The estimated standard error

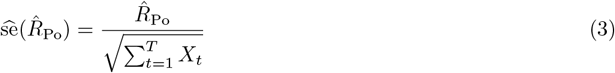

depends on 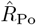 and the total incidence over the estimation window, but not the “noisiness” of the time series. This is a consequence of the equidispersion property of the Poisson distribution, which implies 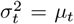, with 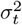 the conditional variance Var(*X*_*t*_ | past). Derivations of equations (2) and (3) are provided in Supplement B.1.1, where we also discuss challenges due to the dynamic nature of model (1).

The estimated standard error (3) can be used to derive Wald-type confidence intervals, which for a confidence level of 95% are given by 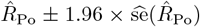. Empirical coverage fractions of such intervals in simulation studies are a straightforward way to assess uncertainty quantification under varying conditions. We will address this in Section 3.1.

### 2.2 Overdispersion and how to account for it

#### 2.2.1 Impact on the Poisson estimator

In practice, infectious disease incidence data usually display conditional overdispersion rather than equidispersion, i.e., 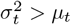. This may be due to a variety of factors, including superspreading events (Lloyd-Smith et al., 2005) and incomplete reporting (Bracher and Held, 2020). The Poisson assumption will then cause our inference for *R* to be overconfident; see Supplementary Section A for an intuitive illustration. Well-known theory on generalized linear regression (Fahrmeir et al., 2009, Sec. 5.5) implies that expression (3) underestimates the true standard error of 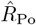 by a factor of 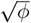, with

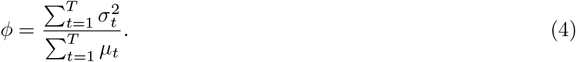

The *variance inflation factor ϕ* depends on the relationship between *µ*_*t*_ and 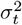. As we shall see in Section 3.2, *ϕ* may be quite large in practice, meaning that standard errors estimated under the Poisson assumption may be considerably too small.

Translating equation (4) to the width of the resulting confidence interval, a (1 − *α*) confidence interval from the Poisson renewal equation model has coverage probability

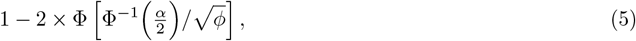

where Φ is the cumulative distribution function (CDF) of the standard normal distribution. This matches the intended level (1 − *α*) only if *ϕ* = 1, and decreases in presence of overdispersion, i.e., if *ϕ >* 1. The details on the derivation of the coverage probability (5) are provided in Supplement B.2.2.

In the following we will outline three options to account for overdispersion, all of which have been applied in the literature. While this is typically motivated by the desire to estimate overdispersion parameters (Ho et al., 2023) and general model realism (Gressani et al., 2022), our focus is on uncertainty estimation.

#### 2.2.2 Quasi-Poisson regression

Quasi-Poisson regression, used by Goldstein et al. (2023), serves to correct the uncertainty quantification for the Poisson model by estimating the variance inflation factor *ϕ* from equation (4). Assuming a linear relationship between *µ*_*t*_ and 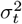, it can be estimated from the residuals of the Poisson model (see Supplement B.2). The corrected standard error is then obtained by multiplying expression (3) with 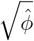. We note that quasi-Poisson regression is not a full probability model, making it unsuitable e.g., for sampling future trajectories.

#### 2.2.3 Negative binomial regression

The most common full probability model to generalize the Poisson assumption is the negative binomial distribution. We here parameterize it by its mean and an overdispersion parameter. The latter quantifies the overdispersion relative to a Poisson distribution (which loosely speaking corresponds to an overdispersion parameter of zero). The overdispersion parameter can be handled in two ways (Cameron and Trivedi, 1998, p.73), which we call *NegBin-L* and *NegBin-Q*, as they imply *linear* and *quadratic* mean variance relationships. Estimates and standard errors are obtained using standard regression methodology and they account for overdispersion.

##### NegBin-L

In the NegBin-L renewal equation

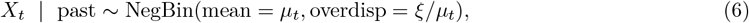

the overdispersion parameter is assumed to be inversely proportional to the mean. This implies a linear mean-variance relationship 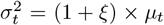 as in the quasi-Poisson model, and can be motivated by overdispersed individual offspring distributions (Ho et al., 2023). While some differences between NegBin-L and quasi-Poisson regression exist (Hilbe, 2011), in our applications we found similar behavior, with point estimates almost identical to those from equation (2).

##### NegBin-Q

We therefore focus primarily on NegBin-Q, defined as

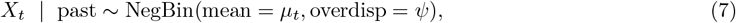

and implying a quadratic mean-variance relationship 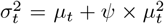. This version, employed e.g., by Held et al. (2005), Gressani et al. (2022) and Brockhaus et al. (2023), can be motivated by variation of *R* over the estimation window. Despite the mean structure *µ*_*t*_ = *R ×* Λ_*t*_ mirroring the Poisson version, the ML estimator 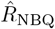 is not identical to 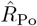 from (2). No closed expression exists, but if overdispersion is sufficiently strong, the approximation

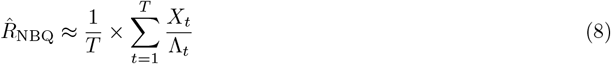

holds. Unlike in (2), the ratios *X*_1_*/*Λ_1_, …, *X*_*T*_ */*Λ_*T*_ thus contribute equally to the estimate. This is because they are all equally informative under a quadratic mean variance relationship, irrespective of the magnitude of Λ_*t*_ and *X*_*t*_ (see discussion in Ver Hoef and Boveng 2007). This also explains why the approximate standard error

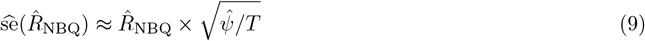

features the number of observations *T* rather than the total incidence as in equation (3). It moreover depends on the estimated overdispersion parameter 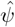, which reflects the noisiness of the time series and can be computed from the model residuals (Supplement B.3). It generally holds that 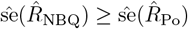, i.e., the estimated standard error is at least equal to its Poisson counterpart. The approximations (8) and (9) are assessed empirically in Supplementary Figure S10.

Maximum likelihood estimation in the negative binomial renewal equation models (6) and (7) can run into convergence issues if the data used for fitting show no or very little overdispersion. This is due to the fact that the overdispersion parameter *ξ* in the NegBin-L model (or *ψ* in the NegBin-Q) is usually estimated on a log-transformed scale to ensure positivity. If the highest likelihood is achieved for a very small value of *ξ*, the likelihood as a function of log(*ξ*) becomes almost flat for log(*ξ*) ≪ 0. This causes numerical failure in the computation of point estimates and/or standard errors. Whenever this occurs, we revert to the Poisson model (1), which in these cases provides a good fit, and report the corresponding point estimates and standard errors.

Quasi-Poisson regression has certain advantages over the negative binomial versions if the mean-variance relationship is mis-specified (Wooldridge, 2010). In our setting, however, both (2) and (8) remain unbiased and consistent in this case, and only their efficiency is reduced. How strongly the two estimators differ is an empirical question we will return to in Section 3.2.

## 3 Results

### 3.1 Simulation study

#### 3.1.1 Setup

To support our claim that Poisson assumptions lead to overconfidence in the estimation of *R* we conduct a simulation study. We combine different distributional assumptions for the data-generating process and the model used for *R* estimation, and compute empirical coverage fractions of Wald confidence intervals. These are given by the fraction of cases in which confidence intervals with a given nominal level contain the true reproductive number *R*.

In the main manuscript, we focus on time series generated from the NegBin-L renewal equation (6), while corresponding results for the Neg-Bin-Q version are available in Supplementary Section C.2.1. We generate 1000 trajectories from each of eight scenarios, which are defined by a combination of three parameters: two values of the reproductive number *R* ∈ {1.5, 2.5}, two levels of overdispersion *ξ* ∈ {1.5, 6 }, and two orders of magnitude, which are controlled by the incidence values used to initialize the simulation. Specifically, the “low magnitude” scenarios begin with sequences of incidence values between 0 and 10, while the “high magnitude” scenarios start at around 100. For each magnitude level, the initialization is fixed, i.e., the same for all low and high-incidence trajectories, respectively. After the initialization, each trajectory was generated using a constant value of *R*, such that in terms of the mean structure the assumptions used for estimation are fulfilled. For all scenarios we used the same shifted gamma serial interval distribution with a mean of 7.5 days and a standard deviation of 2.1 days. This rather long serial interval was chosen to avoid too explosive trajectories, and is motivated by the estimate Vink et al. (2014) provide for respiratory syncytial virus (RSV).

For each simulated trajectory, *R* values were estimated under the four distributional assumptions discussed in Section 2.2. The Poisson and quasi-Poisson models were fitted using the `stats::glm` function in `R` (R Core Team, 2013), while `gamlss:gamlss` (Stasinopoulos and Rigby, 2007) was employed for the negative binomial models. To assess the impact of small sample biases, we applied two different estimation window lengths (7 days and 14 days) across all scenarios. As described in Section 2.2.3, in case of convergence issues due to low degrees of observed overdispersion, we reverted from the negative binomial models to the Poisson version.

#### 3.1.2 Results

Figure 1 shows the sampled trajectories in the different scenarios (left column), along with how often confidence intervals produced under different distributional assumptions cover the true value of *R* (right column). These coverage proportions are shown for the two window lengths (7 and 14 days) and as a function of the nominal coverage level (1 − *α*).

**Figure 1.**
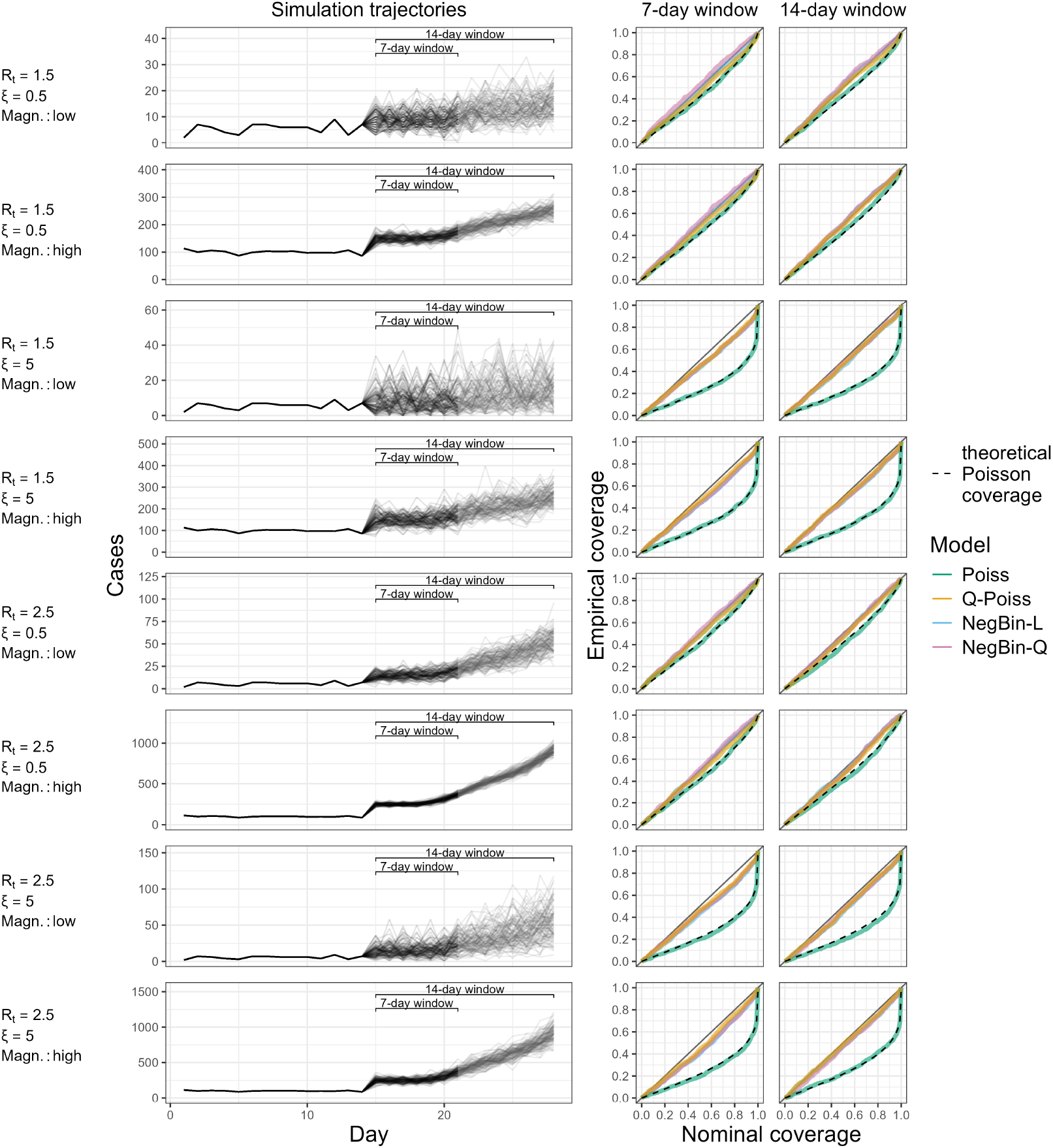
Simulation trajectories generated by the NegBin-L model and empirical coverage of confidence intervals from the four models. The left panel shows 1000 incidence trajectories generated from the renewal equation with the NegBin-L distribution across eight scenarios. These are defined by different parameter combinations specified on the left margin of the figure. The right panels display empirical coverage of the true *R* for Poisson, quasi-Poisson and negative binomial models across nominal coverage levels, using 7-day (middle column) and 14-day (right column) estimation windows. Dashed black lines indicate our theoretical expectation for the coverage of the Poisson model based on equation (5).

The Poisson model shows consistent undercoverage. It is relatively mild in scenarios with low overdispersion (e.g., for *R* = 1.5, *ξ* = 1.5 and low magnitude, the 90% confidence interval covers the true value in 82% of the cases), but becomes quite substantial as overdispersion increases. This is especially evident at high nominal coverage levels. For instance, for *R* = 1.5, *ξ* = 6 and low magnitude, the actual coverage of the Poisson model reaches just 50% for a nominal level of 90%. The observed coverage level of the Poisson model is well-approximated by our theoretical expectation based on formula (5), which is shown as a dashed black line. This underscores the validity of our theoretical arguments.

In contrast, the three models accounting for overdispersion generally exhibit similar coverage behavior with the empirical coverage close to the nominal levels. It thus does not seem crucial whether the mean-variance relationship is specified correctly. A slight tendency to undercover can be spotted for higher nominal coverage levels and larger dispersion values. This issue is less pronounced when the longer 14-day window is used, suggesting the effect of small sample biases when using the 7-day estimation window.

We note that in 18.6% of simulation runs, the fitting algorithm of the negative binomial models did not converge, making it necessary to revert to the Poisson model. In line with our reasoning from Section 2.2.3, this occurred almost exclusively in the scenarios with low overdispersion.

For completeness, we display the empirical distributions of the estimated reproductive number 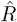 and the associated standard errors in Supplementary Figure S2, as well as the empirical distribution of the overdispersion parameter 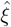, in Supplementary Figure S7. In Supplementary Section C.2.1 we moreover show results for data generated from the NegBin-Q rather than NegBin-L process, which yield qualitatively similar results (again it is not crucial whether the mean-variance relationship is specified correctly, and the NegBin-L and quasi-Poisson models perform reasonably). Lastly, for comparison we show incidence time series from the Poisson model and associated estimates in Section C.2.2. Especially in the high-incidence settings, the resulting bundles of trajectories are very tight, demonstrating that the Poisson distribution in such settings implies an unrealistically low degree of variability. Confidence intervals from both negative-binomial models become somewhat conservative in this setting, a finding we explain in more detail in Section C.2.2 of the Supplement.

### 3.2 Illustration for three outbreaks

To illustrate the impact of the distributional choice in real-world settings, we present three case studies covering different pathogens and locations. These are based on previous applications of `EpiEstim` summarized in Table 1. We use shifted gamma serial interval distributions with means and standard deviations retrieved from the respective references. For the influenza and COVID-19 examples, we also use the corresponding estimation window length. For the Ebola data, we present a variation of the analysis from Green et al. (2022). We aggregate the daily data to weekly totals, rescale the serial interval distribution and employ an estimation window of 4 weeks. This illustrates our point in a weekly setting for a disease with a longer serial interval. As in the simulation study, we show Wald confidence intervals in all cases.

**Table 1.** Summary of the case studies.

| Reference | Disease | Location | Year | Resolution | Serial interval<br>mean (sd) | Estimation<br>window width $T$ |
| --- | --- | --- | --- | --- | --- | --- |
| Nash et al. (2023), data<br>from Riley et al. (2016) | Pandemic<br>influenza | US (military<br>personnel) | 2009–2010 | Daily | 3.6 (1.6) days | 7 days |
| Richter et al. (2022) | COVID-19 | Austria | 2021–2022 | Daily | 3.37 (1.83) days | 13 days |
| Green et al. (2022) | Ebola | Guinea | 2014–2015 | Weekly | 2.19 (1.33) weeks | 4 weeks |

Figure 2 shows the three incidence time series (top row) and *R* estimates based on the different distributional assumptions. The second row compares the Poisson and quasi-Poisson models. While the point estimates agree by definition, the Poisson version (shown in green) yields much narrower 95% confidence intervals. For influenza and COVID-19, they are visually indiscernible from the point estimate. For Ebola, the intervals are wider, as suggested by equation (3) for lower incidence counts. Nonetheless, they rarely include *R* = 1.0, implying high confidence about epidemic growth or decline. The intervals from the quasi-Poisson model (shown in orange), on the other hand, often contain values above and below *R* = 1.0.

**Figure 2.**
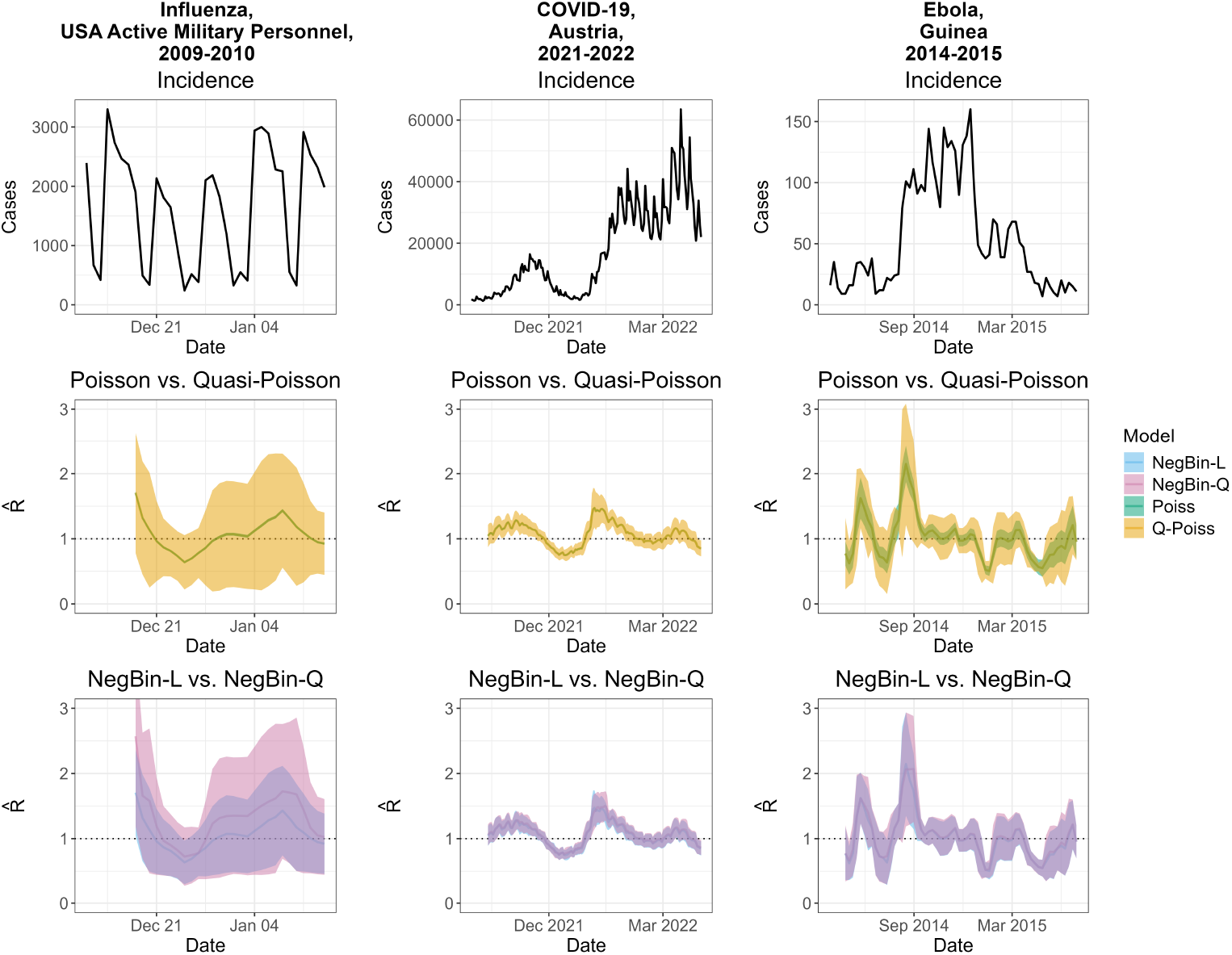
Incidence and estimated effective reproductive numbers for three outbreaks: influenza among active military personnel in the USA, 2009–2010 (left column); COVID-19 in Austria, 2021–2022 (middle); and Ebola in Guinea, 2014–2015 (right). The first row shows case incidences, which are in daily resolution for influenza and COVID-19, and in weekly resolution for Ebola. The second row shows a comparison of the *R* estimates from the Poisson and quasi-Poisson models. All estimates are aligned with the last day of the respective estimation window. The third row shows a comparison of the *R* estimates from the NegBin-Q and NegBin-L models. An additional plot overlaying the quasi-Poisson and NegBin-L versions as well as the estimated overdispersion parameters are available in Supplementary Figures S8 and S9.

The third row overlays results from the NegBin-L model (which behaves similarly to the quasi-Poisson model; Supplementary Figure S9), and the NegBin-Q model. The confidence intervals are comparable for Ebola and COVID-19. For influenza, the NegBin-Q intervals are wider, and there are considerable differences in the point estimates. These are due to the different weighting of the observations in estimators (2) and (8), in conjunction with weekday effects. We discuss this in detail in Supplementary Section A. We note that weekday effects also occur to a lesser degree in the COVID-19 example, but the resulting differences in estimates and confidence intervals are minor.

Ignoring weekday effects can be argued to yield a mis-specified model, and indeed this aspect receives some discussion in Nash et al. (2023). The resulting unexplained variability is absorbed by the overdispersion parameters, leading to confidence intervals which may become overly wide. Despite these issues, we kept the influenza example to illustrate the difference between NegBin-L and NegBin-Q, as well as the difficulty of fitting time-homogeneous models to daily data.

## 4 Discussion

We argued that unjustified Poisson assumptions lead to overconfident estimation of reproductive numbers and discussed options to account for overdispersion. The quasi-Poisson and NegBin-L versions agreed well in terms of point estimates and uncertainty intervals. The NegBin-Q approach yielded somewhat different results, but in simulation studies we found that uncertainty quantification was reliable even if the assumed mean-variance relationship was incorrect. In applications to influenza, COVID-19 and Ebola, all three extensions yielded considerably widened uncertainty intervals relative to the Poisson model, which included the threshold *R* = 1.0 considerably more often. Differences between estimates from the NegBin-L and quasi-Poisson models on the one hand and the NegBin-Q model on the other hand turned out to be substantial in the case study on influenza. This, however, could be attributed to unaccounted weekday effects, meaning that such discrepancies are indicative of more general model mis-specification issues.

In light of these results, we strongly recommend to account for overdispersion in the estimation of reproductive numbers. While the conjugate prior approach from Cori et al. (2013) is difficult to extend, standard regression functions for likelihood-based inference can be adapted. We are aware that an MCMC-based extension of `EpiEstim` to account for overdispersion in a Bayesian framework is under development, and we recommend it to future users.

Alternative tools like `EpiNow2` (Abbott et al., 2020) and `EpiLPS` (Gressani et al., 2022) already account for overdispersion. `EpiNow2` further avoids the need for estimation windows and handles delays between infection events and reporting. This makes estimates more aligned with the actual infection dynamics. Due to its increased complexity, however, `EpiNow2` requires more tuning and computational effort, which may present an obstacle to some users.

We conclude by discussing some limitations of our arguments and suggested extensions. By using off-the-shelf generalized linear models, we conflate different sources of overdispersion, such as superspreading, underreporting, and other reporting artifacts. It is thus difficult to assign any mechanistic interpretation to the estimated overdispersion parameters, and they cannot serve to derive implications on disease emergence and control (Lloyd-Smith et al., 2005). Moreover, in the examples of influenza in US military personnel and (to a lesser degree) COVID-19 in Austria, the model appeared to be mis-specified due to weekday patterns. These may inflate the estimated overdispersion parameters and standard errors. More generally, estimating the overdispersion parameter along with *R* from short time windows may lead to instabilities. For negative binomial regression, it is known that small-sample biases can occur (Kenne Pagui et al., 2022), with a tendency to underestimate standard errors. Lastly, many other layers of complexity like unobserved initial generations in an outbreak (Brizzi et al., 2022) and changes in testing volumes (Goldstein et al., 2023) could be addressed. However, we consider these outside the scope of the present article.

A promising regression-based approach replacing sliding window approach by splines has recently been proposed by Nouvellet (2025), and can remedy some of the aforementioned weaknesses. It estimates the temporal variation in *R* flexibly, potentially accounting for covariates like weekday effects. Unfortunately, the new framework currently only supports the Poisson distribution, and is thus likewise affected by the pitfalls we described. Extensions, however, could easily be implemented, potentially even allowing for time-varying overdispersion parameters via the `gamlss` framework (Stasinopoulos and Rigby, 2007).

## Data Availability

In this study, only data previously available online were used. All data are available at

https://github.com/barbora-sobolova/epiestim_overdispesion

## Reproducibility

The repository https://github.com/barbora-sobolova/epiestim_overdispesion/ contains code and data to reproduce our analyses.

## A Intuitive illustration of maximum likelihood estimation

To strengthen intuition, we provide a graphical illustration of likelihood inference under different models. The top panel of Figure S1 shows a 7-day window (3–9 January 2010) taken from the US influenza data discussed in Section 3.2. The incidence is overlaid with Λ_*t*_ from equation (1). The remaining panels show the log-likelihood contributions of the seven individual observations (grey) and the overall log-likelihood functions (black) for the Poisson and negative binomial models, all on a relative scale. For the negative binomial versions, the overdispersion parameters are fixed at the maximum likelihood estimates.

**Figure S1.**
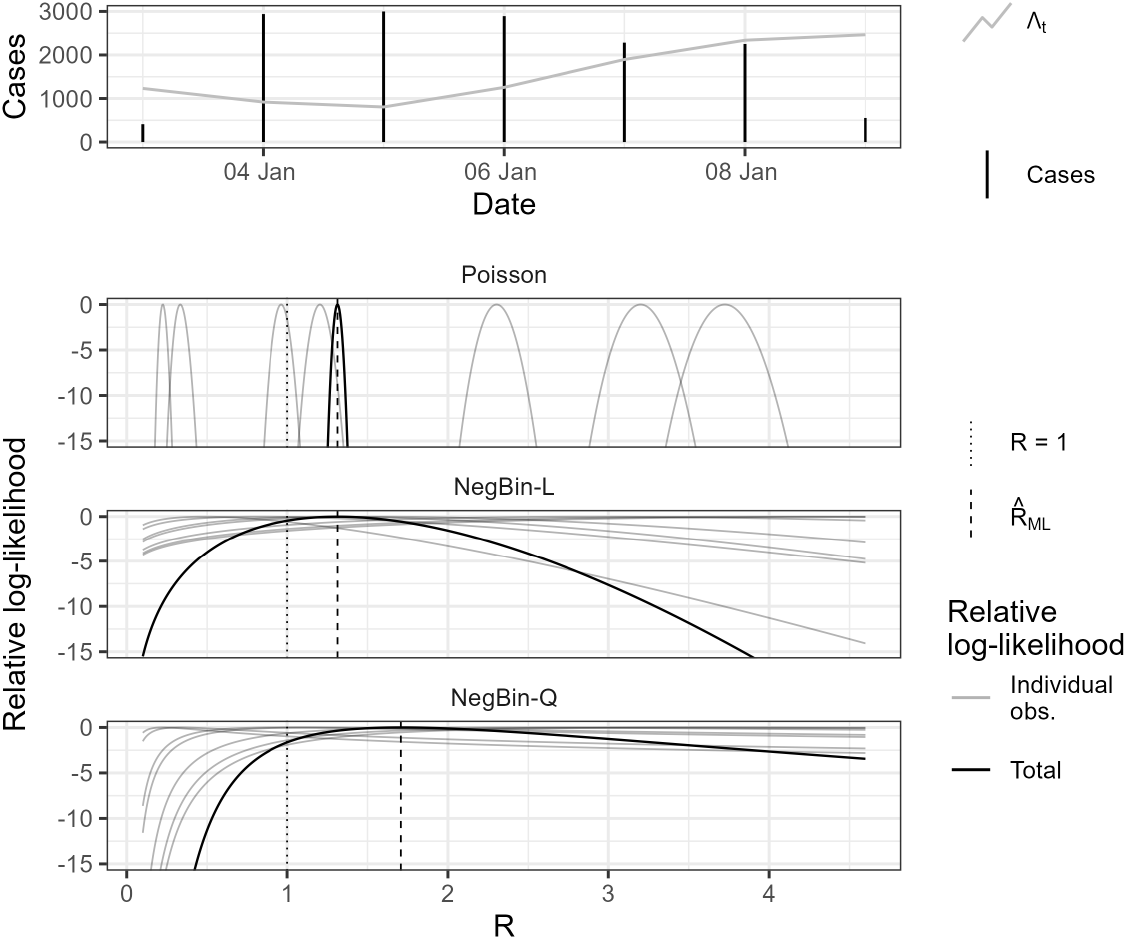
Illustration of maximum likelihood estimation in the Poisson, NegBin-L and NegBin-Q models. In the three bottom panels, light grey lines show the individual likelihood contributions, while black lines show the overall likelihood functions (i.e., the sums of the respective contributions). All of these are shown on a relative scale such that their peak value is zero. Maximum likelihood estimates are highlighted by vertical dashed lines, while the threshold value *R* = 1.0 is marked by a dotted line.

Due to the equidispersion assumption and high observed counts, the Poisson log-likelihood contribution functions are narrow, and the overall log-likelihood function exhibits a sharp peak. The curves for the negative binomial models are flatter as the model adapts to the higher degree of dispersion. Since 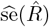 reflects the inverse curvature of the log-likelihood function at its peak (see Supplement B.1.1), it is clear that the Poisson model issues much more confident estimates. Notably, the Poisson likelihood clearly favors a value of *R >* 1, while for the negative binomial models, the relative likelihood is rather high for *R* = 1.

We moreover notice a shift in the point estimates from different models (dashed vertical lines). While they are almost identical for the Poisson and NegBin-L models 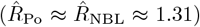, we obtain a higher value for NegBin-Q (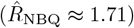). This is a result of the particular alignment of Λ_*t*_ and *X*_*t*_, caused by weekday effects, and the different weights the *T* = 7 observations receive in the two schemes. To understand this, note that both estimators are weighted averages of the daily ratios *X*_*t*_*/*Λ_*t*_, *t* = 1, …, *T* . For the NegBin-Q model, this average is unweighted as in equation (8). For the Poisson model, equation (2) can be written as

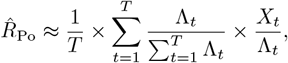

and also applies in good approximation for the NegBin-L model (McCullagh and Nelder, 1989, p. 199). Here, the ratios *X*_*t*_*/*Λ_*t*_ thus enter with weights proportional to Λ_*t*_. As seen in the top panel of Figure S1, the highest values of *X*_*t*_ occur for the lowest values of Λ_*t*_. These differences in weighting thus explain why the NegBin-Q estimate is considerably higher than its counterparts from the Poisson and NegBin-L models.

## B Mathematical details

### B.1 Details on the Poisson model, equation (1) in the main text

#### B.1.1 Maximum likelihood estimation (equations (2) and (3) in the main text)

Here we derive the formulas (2) and (3) using standard maximum likelihood theory. In the following we denote the vector of observed values in the estimation window by **X** = (*X*_1_, …, *X*_*T*_) and substitute *µ*_*t*_ = *R*Λ_*t*_. Due to the *D*-th order Markov structure of the Poisson renewal equation model (1), the log-likelihood is given by a sum of *T* Poisson log-likelihoods,

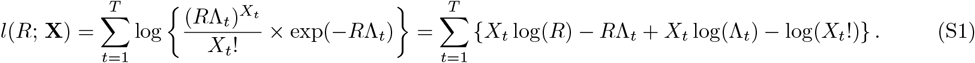

Taking the derivative with respect to *R* we obtain the score function

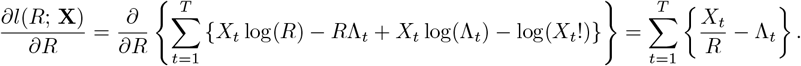

Next we set the score function equal to zero and solve for *R*,

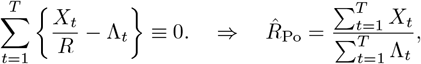

which results in the maximum likelihood estimator. In order to find an estimate of its variance, we compute the observed Fisher information, which involves taking the second derivative of the log-likelihood function,

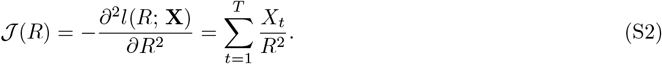

We then obtain the formula of the standard error of 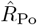 by inverting the observed Fisher information at the maximum likelihood estimate and taking the square root,

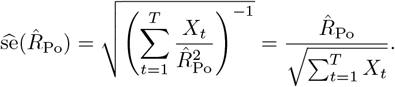

We note that unlike in a classic generalized linear regression model (GLM), the “covariate” Λ_*t*_ is itself random, and all Λ_*t*_, *X*_*t*_, *t* = 1, …, *T* are dependent. This invalidates some of the basic assumptions underlying maximum likelihood theory for GLMs. Due to the Markov structure of the process (1), however, the log-likelihood function (S1) and the observed Fisher information (S2) are nonetheless correct, and the `glm` implementation in R can be used to evaluate them. It is non-trivial to show that maximum likelihood estimators preserve their usual properties in stochastic process models like (1), as relevant mixing properties need to be established. This has been done in related stationary models from the INGARCH family (see e.g., Fokianos et al. 2009), but in practice maximum likelihood estimators and associated standard errors are widely used and have been found to be reliable even when their properties have not been formally demonstrated (see e.g., Paul et al. 2008). For model (1), an additional challenge arises from the fact that the process is non-stationary and potentially explosive. Mixing properties in related settings have been explored by Doukhan et al. (2022), but we consider these aspects outside the scope of the present applied article.

#### B.1.2 Comparison to Bayesian estimation

In the `EpiEstim` package, Bayesian estimation of *R* in the model (1) is based on a conjugate gamma prior distribution. Specifying the prior distribution as

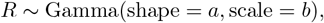

the posterior distribution given the data **X** = (*X*_1_, …, *X*_*T*_) is likewise a gamma distribution. Specifically, we obtain (Cori et al., 2013, Supplementary Material, page 3)

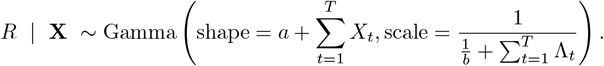

The first two posterior moments of *R* are consequently given by

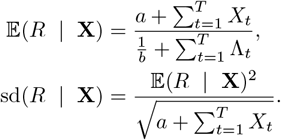

This strongly resembles the maximum likelihood estimator 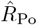 from (2) and its estimated standard deviation given in equation (3). Indeed, the Bayesian scheme can be seen as a regularized maximum likelihood estimate resulting from the addition of a pseudo observation with Λ_0_ = 1*/b* and *X*_0_ = *a*. The default values in `EpiEstim` are *a* = 1, *b* = 1*/*5, meaning that for moderately high incidence values they have little influence on the result.

### B.2 Details on the quasi-Poisson model

#### B.2.1 Estimation

The most common way to estimate the overdispersion parameter 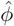, implemented in the `R` function `glm(…, family = quasipoisson)`, is

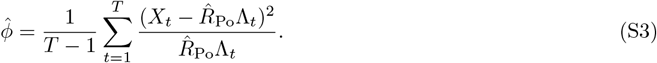

This is simply the empirical variance of the Pearson residuals in the Poisson model. The standard errors in expression (3) are then corrected to

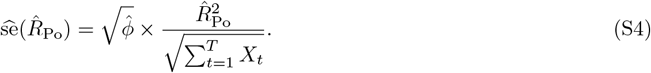

#### B.2.2 Implications for confidence intervals

For *ϕ* fixed, the (1 − *γ*) Wald confidence interval of the quasi-Poisson model becomes

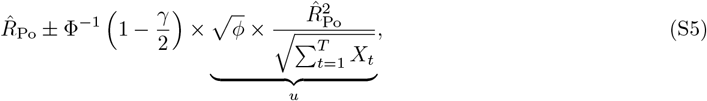

where Φ(·) denotes the cumulative probability function of the standard normal distribution and Φ^−1^(·) its quantile function.

If the assumptions of the quasi-Poisson regression are fulfilled, we can use this to derive the expected coverage level of uncorrected Poisson-based confidence intervals. To this end, note that the quasi-Poisson interval at level (1 − *γ*) is constructed such that

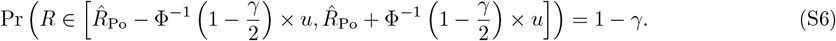

Now setting

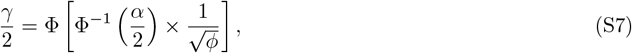

it is easy to show that the (1 − *γ*) confidence interval of the quasi-Poisson model and the (1 − *α*) interval of the Poisson are identical. This is because the relationship between the (1 − *γ/*2) and (1 − *α/*2) quantiles of the standard normal distribution is as follows

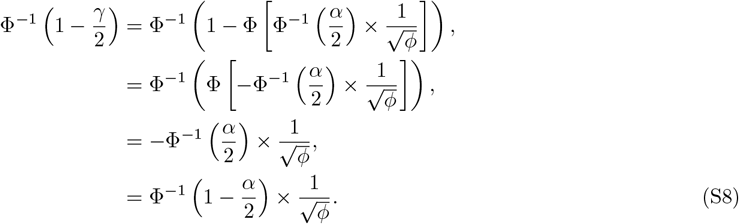

Substituting for Φ^−1^(1 − *γ/*2) in (S5) we get

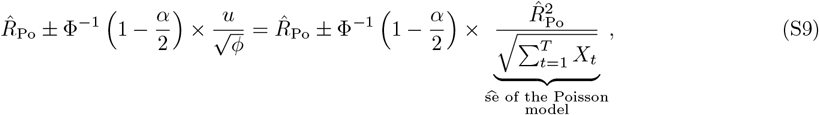

which is the formula of the (1 − *α*) Wald confidence interval of the Poisson model. Now, constructing this (1 − *α*) Poisson confidence interval, while the quasi-Poisson model holds, corresponds to plugging (S8) into (S6), which yields the expected coverage

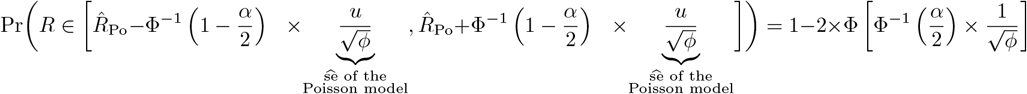

of the Poisson-based confidence interval.

### B.3 Details on the NegBin-Q model, equation (7) in the main text

#### B.3.1 Maximum likelihood estimation, equations (8) and (9) in the main text

The negative binomial distribution with mean *µ* and dispersion parameter *ψ* has probability mass function

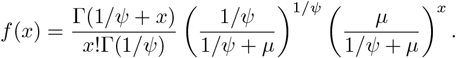

Following the same logic as in Supplement B.1.1, the log-likelihood function of the NegBin-Q model (7) is

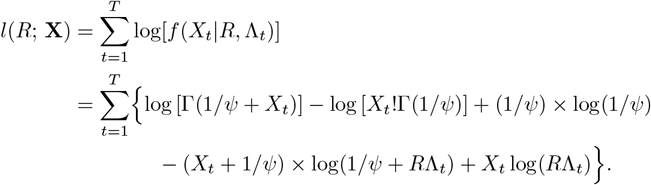

Now we differentiate with respect to *R* to obtain the score equation

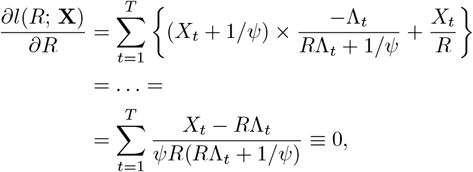

which is equivalent to

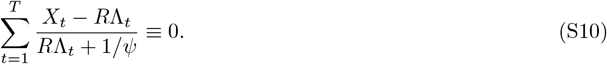

We now assume that there is substantial overdispersion, which we formalize as

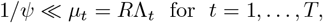

implying that

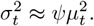

Equation (S10) then simplifies to the estimator from equation (8) as

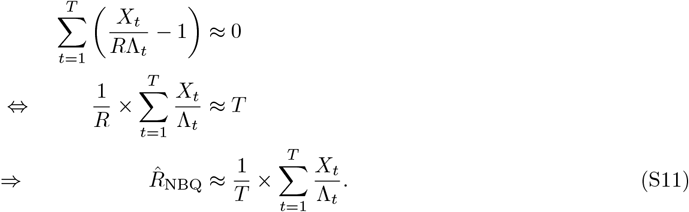

Next we again compute the observed Fisher information with respect to *R*,

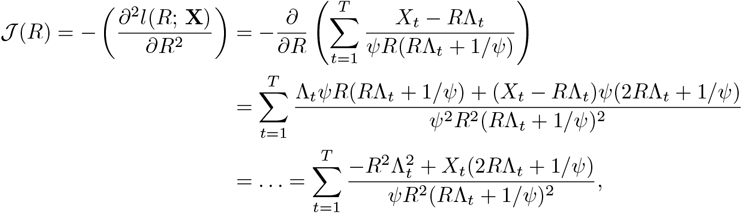

Now again assuming that 1*/ψ* ≪ *µ*_*t*_ = *R*Λ_*t*_ this simplifies to

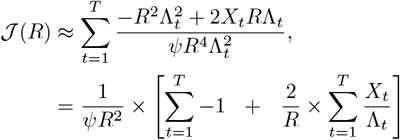

Now evaluating this at the (approximate) maximum likelihood estimator (S11) we obtain

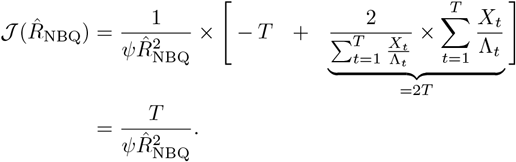

As before, the standard error of 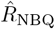 then results as

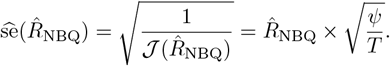

#### B.3.2 Estimation of the overdispersion parameter *ψ*

The overdispersion parameter *ψ* can be estimated as

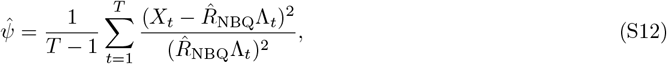

which resembles equation (S3), but is different due to the square in the denominator. This estimator, which to our knowledge has not been proposed previously, is motivated by the fact that under model (7) we have

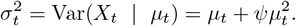

Under our additional assumption *ψ* ≫ 1*/µ*_*t*_ this simplifies to

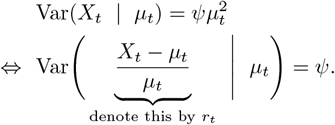

Now note that we moreover have

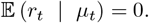

Due to the Markov property of {*X*_*t*_}, the *r*_1_, …, *r*_*T*_ are hence independent random variables with mean zero and identical variance *ψ*. The empirical variance (S12) of *r*_1_, …, *r*_*T*_ is thus a natural estimator of *ψ*.

## C Details on the simulation study

### C.1 Empirical distribution of the 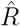 estimates

Supplementary Figure S2 shows the empirical distribution of the 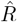 estimates across the different scenarios and 1000 simulation runs each. The point estimates follow similar distributions across all four models and scenarios. In contrast, clear differences can be spotted in the distribution of estimated standard errors. The Poisson model produces consistently low estimated standard errors, whereas models accounting for overdispersion yield a broader range of higher values.

**Figure S2.**
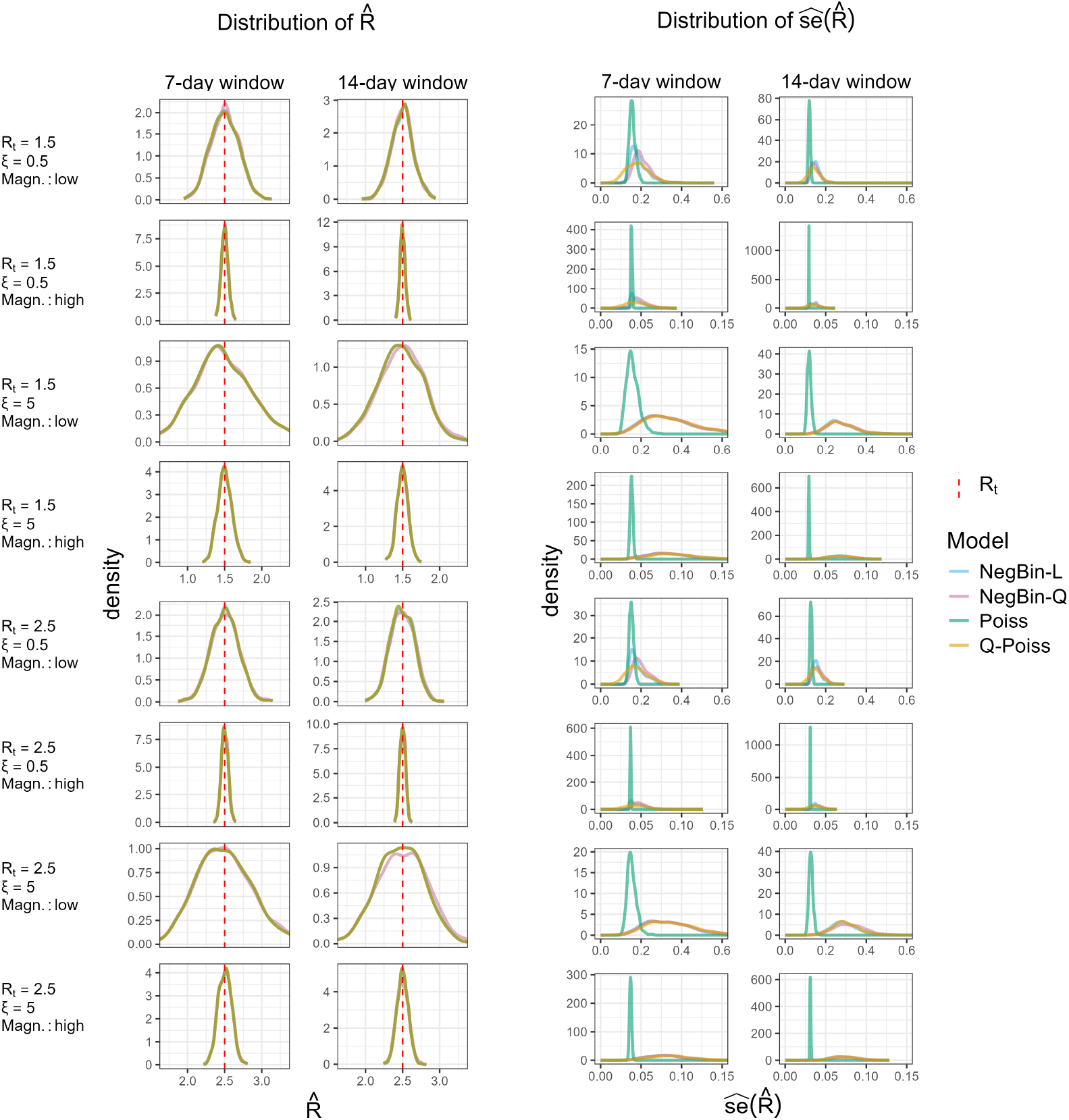
Empirical distribution of the 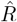 estimates (two left columns) and their standard errors (two right columns) under the NegBin-L distributional assumptions. Different parameter combinations are specified on the left margin of the figure. The red dashed line shows the true value of the effective reproductive number *R*.

### C.2 Supplementary simulation scenarios

#### C.2.1 NegBin-Q simulation scenarios

We further simulated eight scenarios assuming the NegBin-Q distribution as the true data-generating process, instead of NegBin-L. Similarly to the simulation in the main text, approximately 14% of simulation runs in low dispersion settings did not converge, requiring to revert to the Poisson model for these particular runs. The empirical coverage results, shown in Supplementary Figure S3, are overall similar to those from Figure 1. Again we see strong under-coverage of the Poisson model, and slight under-coverage of the other models in settings with high dispersion and short estimation windows. A difference lies in the fact the severity of Poisson undercoverage also increases with higher incidence magnitude. This is a direct consequence of the quadratic mean-variance relationship of NegBin-Q process used to generate the data. We note that no explicit formula for the actual coverage of the Poisson model is available in this setting as (unlike in the NegBin-L case) equation (5) cannot be simplified in a suitable manner.

**Figure S3.**
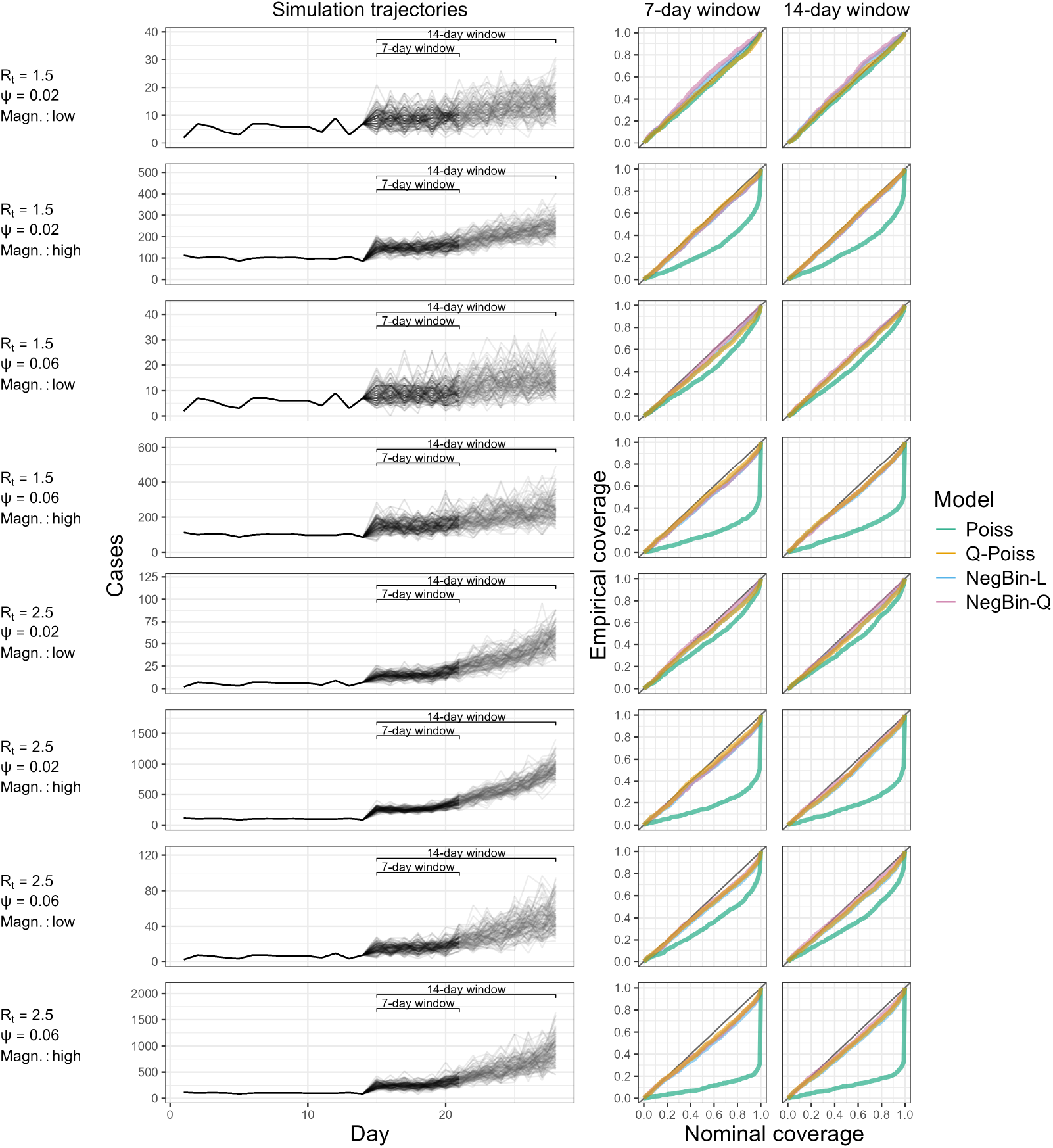
Simulation trajectories generated by the NegBin-Q model and empirical coverage of the four models. The left panel shows 1000 incidence trajectories generated from the renewal equation with the NegBin-Q distribution across eight scenarios defined by different parameter combinations specified on the left margin of the figure. The right panels display empirical coverage of the true *R* for Poisson, quasi-Poisson and negative binomial models across nominal coverage levels, using 7-day (middle column) and 14-day (right column) estimation windows.

The empirical distribution of 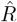 estimates under NegBin-Q, shown in Supplementary Figure S4, is similar to that observed when NegBin-L is the underlying count distribution. Point estimates are comparable across all models, while the standard errors from the Poisson model are located around smaller values than those from the overdispersed models.

**Figure S4.**
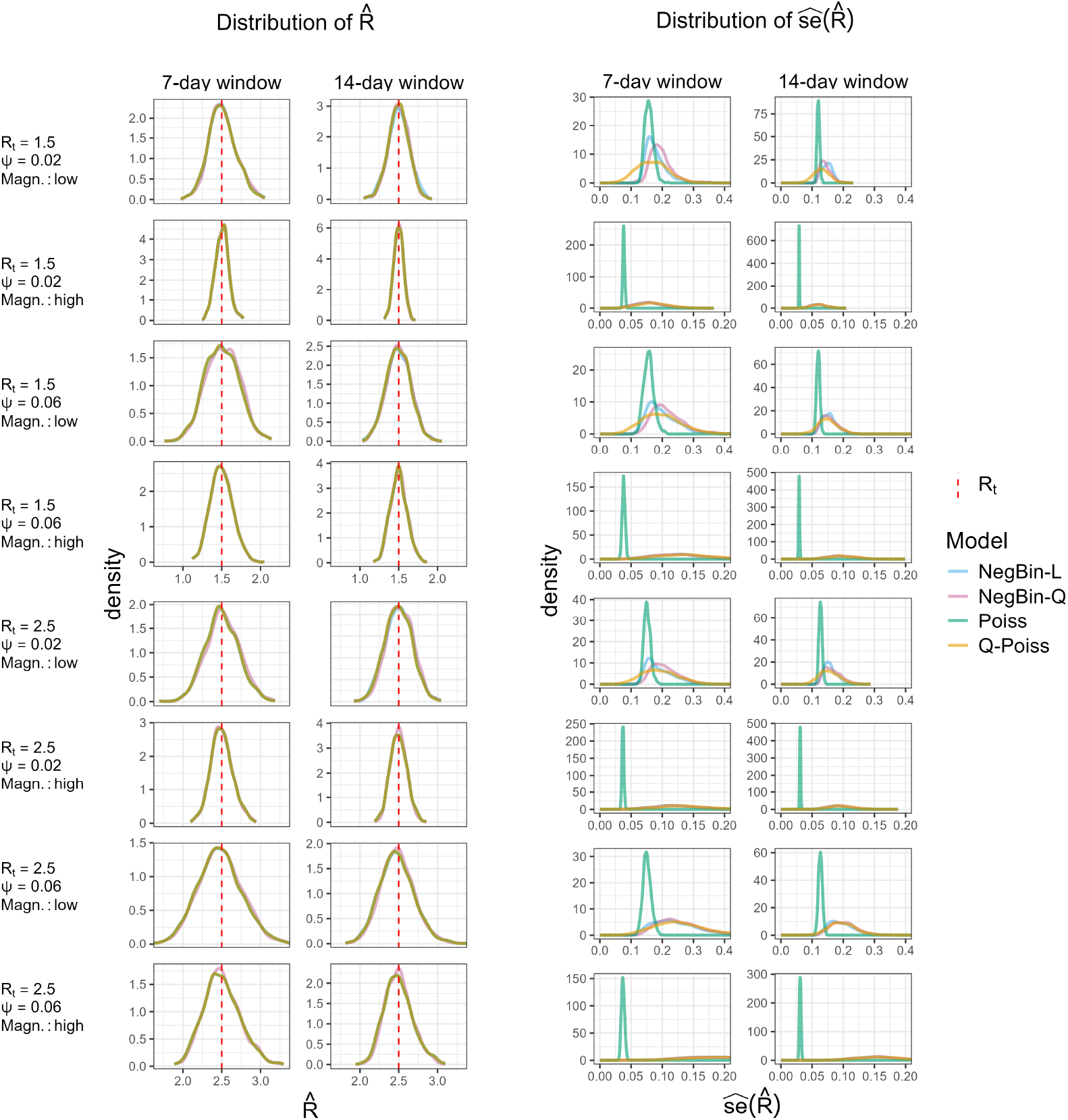
Empirical distribution of the 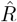 estimates (two left columns) and their standard errors (two right columns) under the NegBin-Q distributional assumptions. Different parameter combinations are specified on the left margin of the figure. The red dashed line shows the true value of the effective reproductive number *R*.

#### C.2.2 Poisson simulation scenarios

To explore the properties of overdispersed models in settings with no overdispersion present, we conducted four additional simulation scenarios. The parameters were identical to those in Section 3.1, except that there is no dispersion parameter to vary. Due to the absence of overdispersion, the fitting algorithm of the negative binomial models managed to converge only in roughly one third of the simulation runs. Supplementary Figure S5 displays the generated trajectories and corresponding coverage results. As expected, the empirical and the nominal coverage are aligned best for the true Poisson model. For the quasi-Poisson model, we recognize weak tendencies to undercover. Since the tendency is stronger for the short estimation window, we ascribe this to the effect of small-sample biases. In contrast, the negative binomial models seem to reach higher empirical coverage than the specified nominal levels. This can be explained by the inability of the negative binomial models to handle underdispersion. They detect occasional overdispersed trajectories, while for underdispersed ones, the Poisson estimate is used, which can be seen as a limiting case for the overdispersion getting vanishingly small. As a result, confidence intervals from the negative binomial models are, on average, somewhat wider.

**Figure S5.**
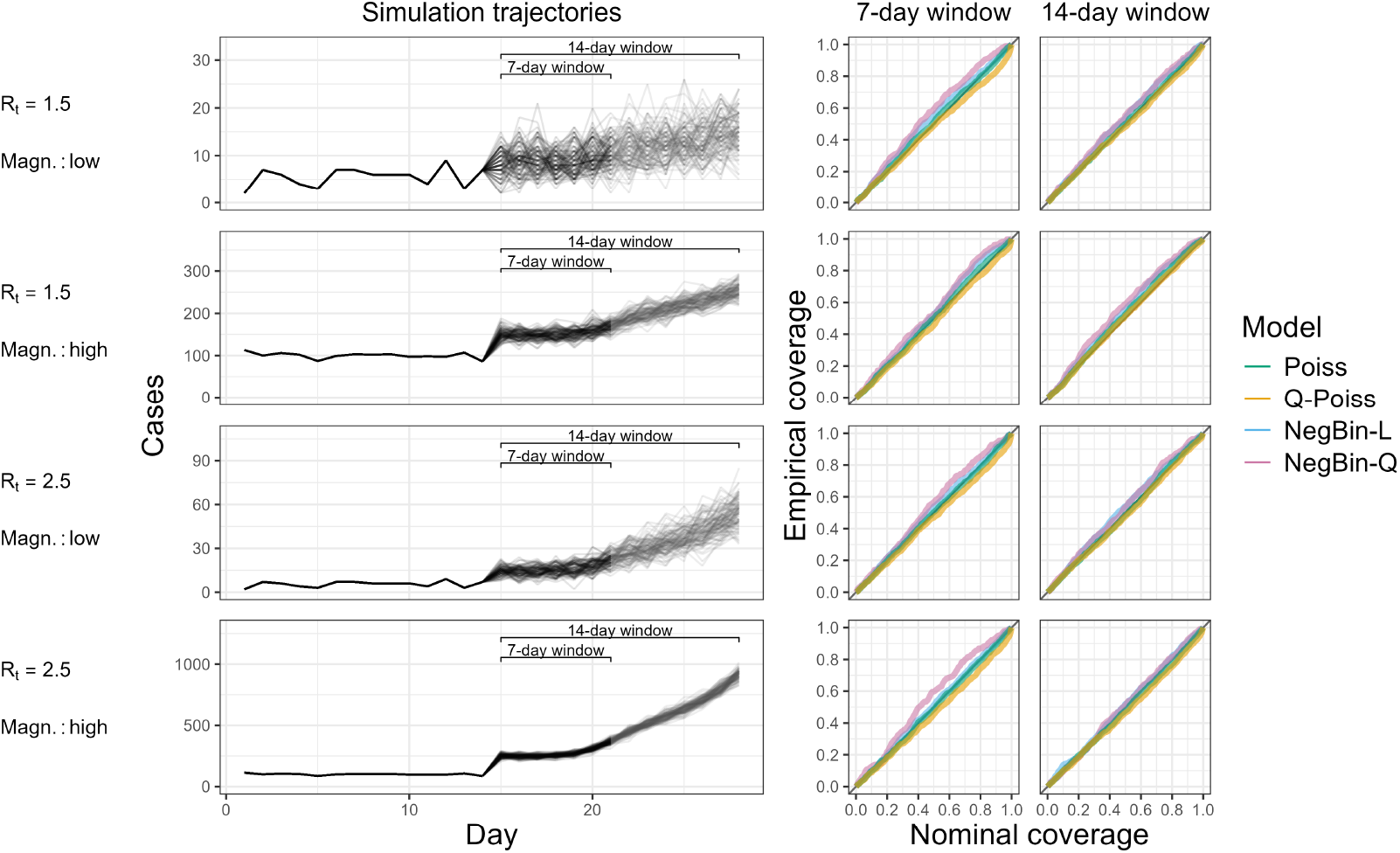
Simulation trajectories generated by the Poisson model and empirical coverage of the four models. The left panel shows 1000 incidence trajectories generated from the renewal equation with the Poisson distribution across four scenarios defined by different parameter combinations specified on the left margin of the figure. The right panels display empirical coverage of the true *R* for Poisson, quasi-Poisson and negative binomial models across nominal coverage levels, using 7-day (middle column) and 14-day (right column) estimation windows.

### C.3 Empirical distribution of 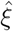 **and** 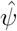

Supplementary Figure S2 shows the empirical distribution of the overdispersion parameter estimates across the different scenarios and 1000 simulation runs each. The estimates in the left column correspond to scenarios discussed in the main text, where NegBin-L is the true data-generating process. We can directly compare the values of the NegBin-L overdispersion parameter *ξ* and quasi-Poisson overdispersion parameter *ϕ* if we shift the quasi-Poisson estimates to the left by one unit. We can notice that for scenarios with low overdispersion, the quasi-Poisson model sometimes suggests values of *ξ <* 1, which corresponds to the simulated trajectories occasionally exhibiting underdispersion. For scenarios with higher dispersion, the estimates are fairly similar. Overall, the estimates are concentrated around the true value with the estimates using the short estimation window (dotted lines) being less accurate.

The right column correspond to the scenarios described in Section C.2.1 with NegBin-Q being the underlying true distribution. Again, the estimates using only the short estimation window seem to be less accurate.

**Figure S6.**
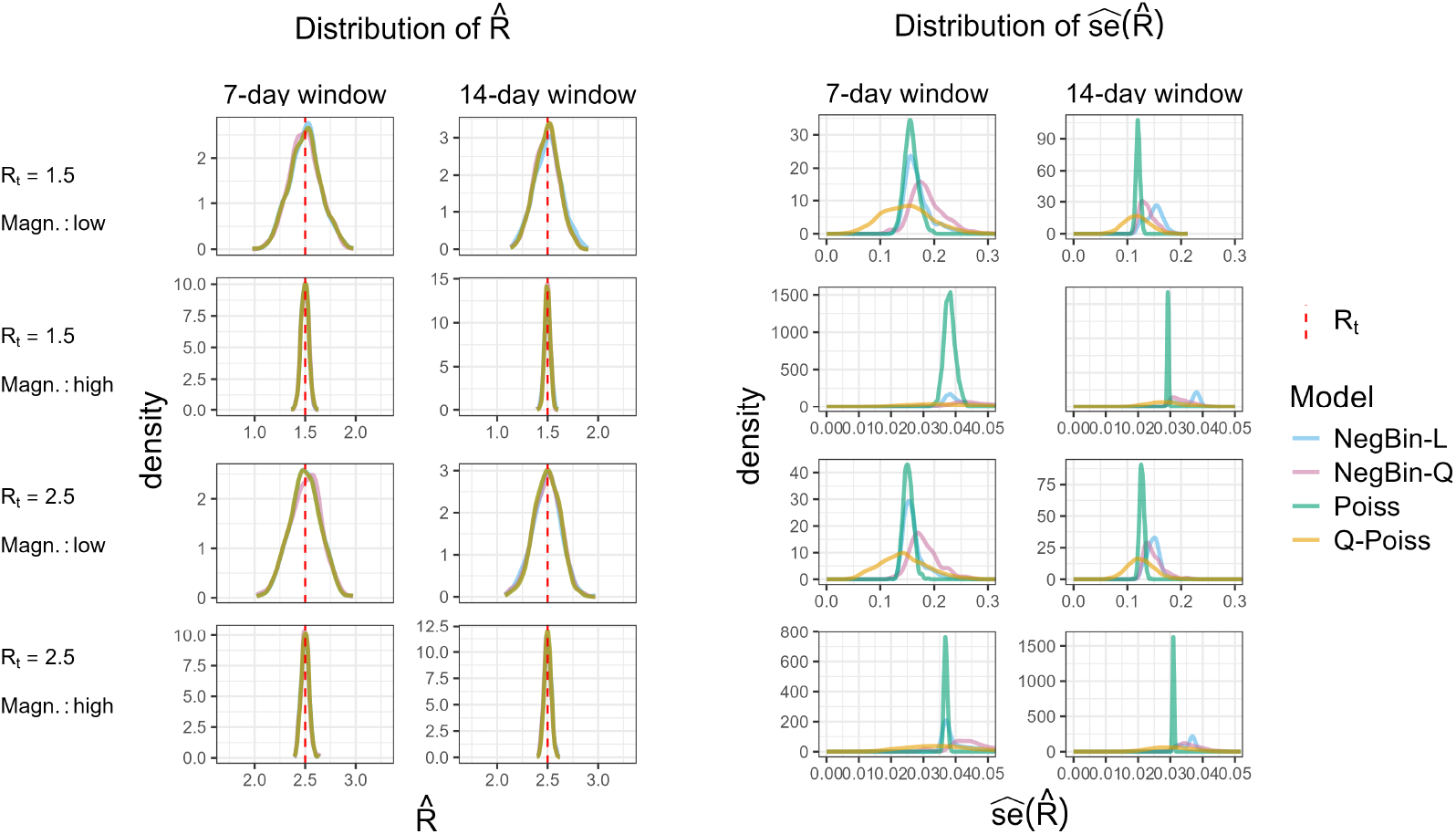
Empirical distribution of the 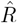 estimates (two left columns) and their standard errors (two right columns) under the Poisson distributional assumptions. Different parameter combinations are specified on the left margin of the figure. The red dashed line shows the true value of the effective reproductive number *R*.

**Figure S7.**
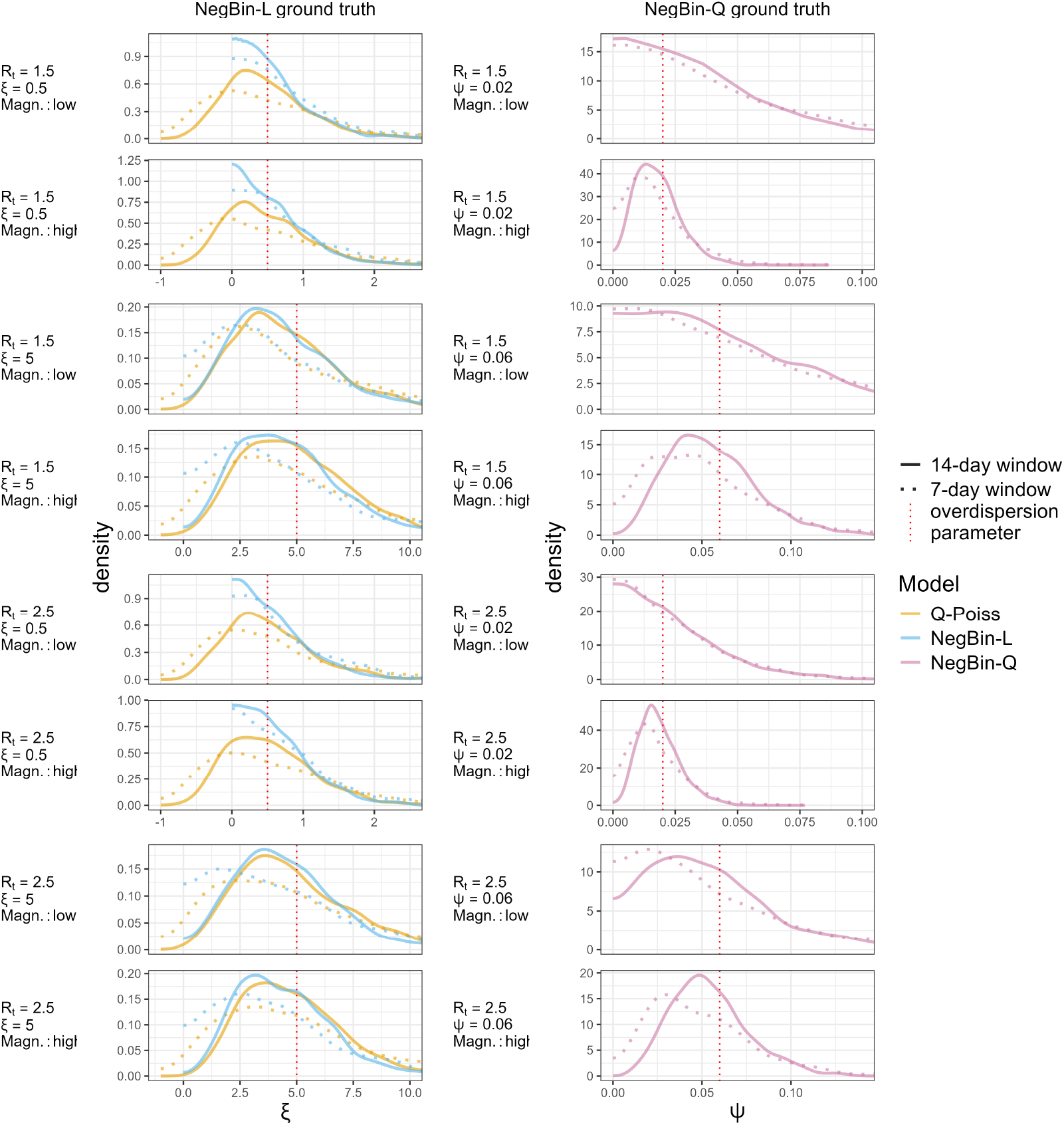
Empirical distribution of the 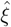 estimates under the NegBin-L distributional assumptions (left column) and 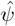 estimates under the NegBin-Q distributional assumptions (right column). Different parameter combinations are specified on the left margin of each panel. The red dashed line shows the true value of the overdispersion parameter.

## D Details on case studies

**Figure S8.**
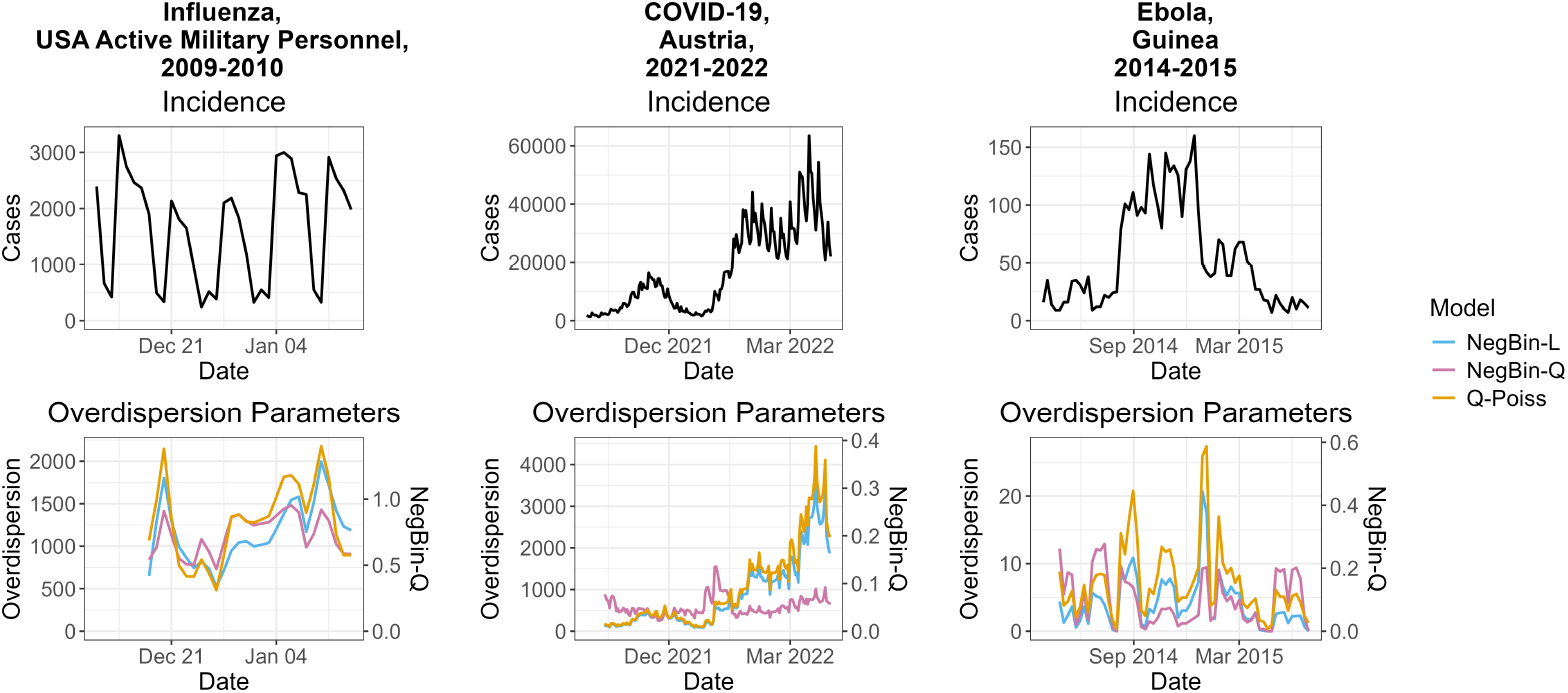
Estimated overdispersion parameters in the three case studies from Figure 2. The left vertical axis refers to the quasi-Poisson and NegBin-L models, while the right vertical axis refers to the NegBin-Q model.

**Figure S9.**
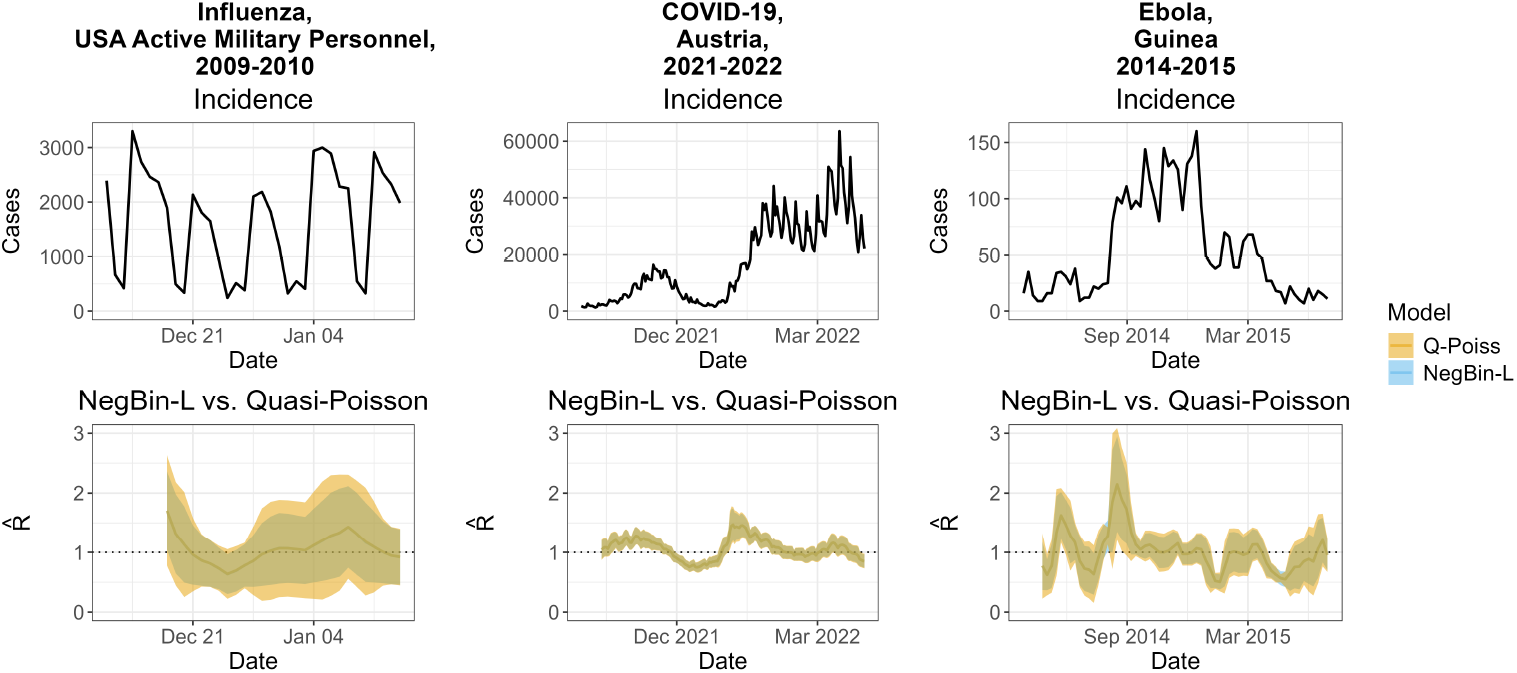
Comparison of R estimates from the quasi-Poisson and NegBin-L models in the case studies from Figure 2.

**Figure S10.**
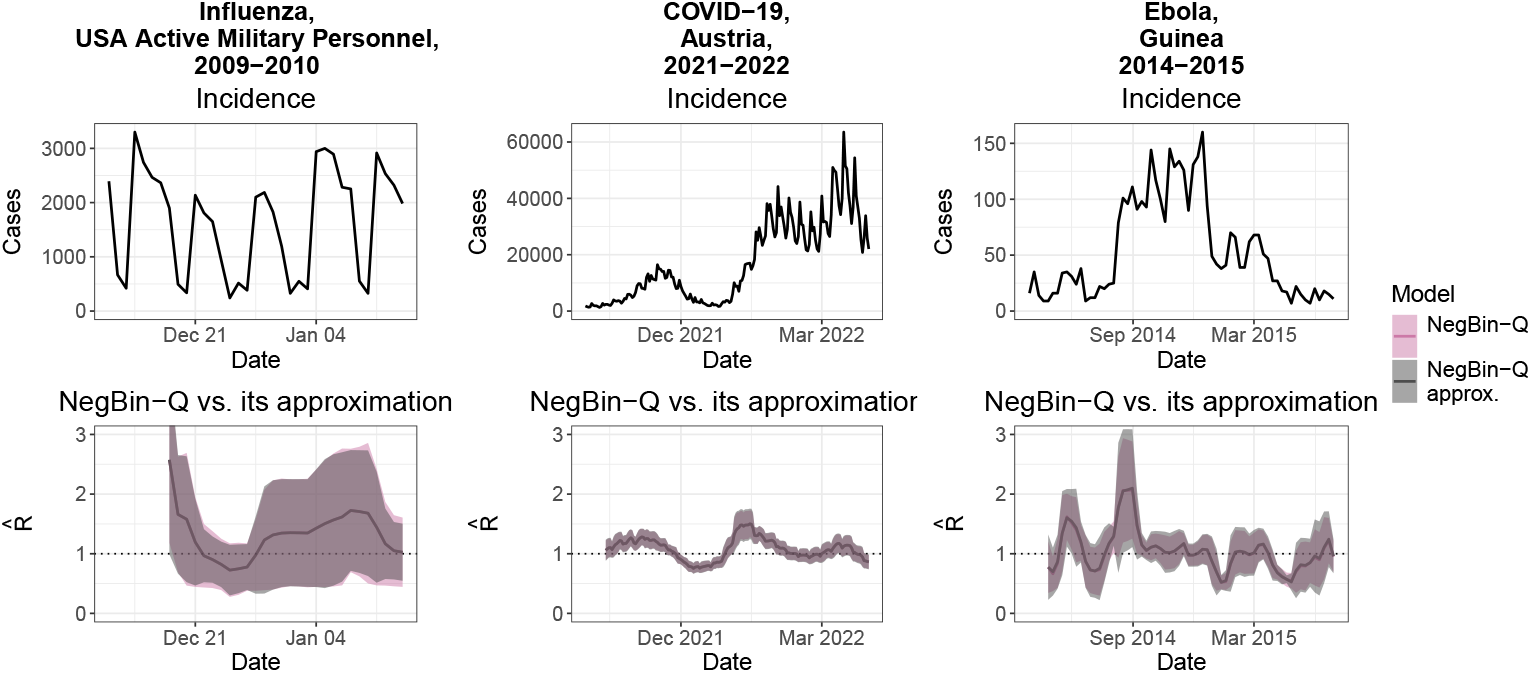
Comparison of *R* estimates and associated Wald confidence intervals from the NegBin-Q model obtained using the `gamlss` package and approximate formulas (8), (9) and (S12). Agreement is overall close, with minor discrepancies for the Ebola case study.

